# Concerns about disease management and psychological stress in SAPHO patients during the COVID-19 epidemic

**DOI:** 10.1101/2020.05.07.20084087

**Authors:** Shuo Zhang, Xinyu Lu, Yihan Cao, Yueting Li, Chen Li, Wen Zhang

**Affiliations:** Department of Rheumatology, Peking Union Medical College Hospital, Peking Union Medical College and Chinese Academy of Medical Sciences, National Clinical Research Center for Dermatologic and Immunologic Diseases, Ministry of Science & Technology, Key Laboratory of Rheumatology and Clinical Immunology, Ministry of Education, Beijing, China, 100730; Institute of Clinical Medicine, Chinese Academy of Medical Sciences & Peking Union Medical College, Beijing, China; Department of Traditional Chinese Medicine, Peking Union Medical College Hospital, Peking Union Medical College and Chinese Academy of Medical Sciences, Beijing, China, 100730

**Keywords:** SAPHO syndrome, COVID-19, psychological condition, disease activity, cross-sectional study

## Abstract

**Objectives:** The coronavirus disease 2019 (COVID-19) epidemic brings potentially impact on the care of patients with rheumatic diseases, including SAPHO syndrome. We aimed to investigate the disease status, concerns about management, and psychological stress in SAPHO patients during the COVID-19 epidemic.

**Method:** A structured questionnaire was distributed online to patients with SAPHO syndrome enrolled in a Chinese cohort study on March 3^rd^, 2020. Patients were ask about the current treatments, disease status, and concerns about disease management during the epidemic. Psychologic stress (scored from 0 to 10 points) and psychological problems were reported by the patients.

**Results:** A total of 157 patients (mean age 38.4 ± 12.3 years, 66.9% females) were included in the study. None of the patients were diagnosed with COVID-19. Sixty-five (41.4%) patients worried about their disease conditions during the epidemic with concerns including medication shortage (73.8%), delay of consultation (46.2%), and disease aggravation (61.5%). Sixty-seven (42.7%) patients had medication withdrawal or dose reduction due to lack of drugs, irregular daily schedule or subjective reasons. The most common psychological problems reported was little interest or pleasure in doing things (66.2%). Patients with progressive disease condition were more distressed and disturbed by the epidemic. Patients with nail involvement felt more worried about their disease conditions than patients without (59.6% vs 31.0%, p =0. 001).

**Conclusions:** The COVID-19 epidemic imposes a negative impact on the disease management and psychological stress in SAPHO patients. Patients’ access to specialty care and medication well as mental stress is of great concern.

## Introduction

The coronavirus disease 2019 (COVID-19) is caused by severe acute respiratory syndrome coronavirus 2 (SARS-CoV-2).^1^ The disease is predominantly found in patients with median age of 44 years old, with around 50% of male cases.^2^ It has spread throughout worldwide by human-to-human transmission via droplets or direct contact since December, 2019.^3^ To interrupt COVID-19 transmission, stringent containment measures are taken widely in China, including isolation of cases, quarantine of contacts, and strict restrictions on personal movement. As a public health emergency of international concern, the COVID-19 epidemic brings about great psychological pressure to the public and may lead to various mental problems. The limited knowledge of COVID-19 and a number of related news reports may aggravate anxiety and fear of healthy people and those who originally had psychological problems.^4^

In this condition, attention should be paid on potentially vulnerable patients suffering from autoimmune diseases due to immuonospression state caused by therapy and requirement of long-term elaborate management. Synovitis, acne, pustulosis, hyperostosis, and osteitis (SAPHO) syndrome is a rare chronic autoimmune disease characterized by osteoarticular and cutaneous manifestations.^5^ Owing to the high rate of false and/or delayed diagnoses and non-ideal therapeutic effect, patients have to endure long-lasting bone and joint pain, which causes increased physical and psychological pressure as well as declining quality of life. Previous studies in in patients with rheumatic diseases revealed that psychological stress induced exacerbation of disease activity.^6,7^ Thus, mental health and quality of life of SAPHO syndrome require attention. Meanwhile, stringent containment measures during the COVID-19 epidemic also bring great challenges in chronic disease management. Accoding to a survey in inflammatory bowel disease, the scheduled follw-up of 70.4% of patients were affected by COVID-19 pandemic.^8^ Thus, it is crucial to clarifying the effects of COVID-19 on patients with SAPHO syndrome.

In this study, we aimed to understand the disease, life, and psychological situations of Chinese SAPHO patients in the COVID-19 epidemic, and to analyze the effect of the COVID-19 epidemic on patients’ psychology.

## Methods

### Participants

We recruited patients who had at least one follow-up between July, 2019 and Jan 2020 in our SAPHO syndrome cohort study.^9^ Participation in the study was voluntarily, and written informed consent was obtained from each patient. Questionnaires were distributed digitally on March 3^rd^, 2020, and were collected on March 5^th^, 2020. An online orientation explanation was provided to each participate before the questionnaire survey. This study complied with the Declaration of Helsinki and was approved by the ethics committee of Peking Union Medical College Hospital (S-K1171).

### Questionnaires

We collected demographic data, disease duration, current treatment, clinical manifestations, and the visual analogue scale (VAS) scores of bone and joint pain. The levels of psychologic stress, which were scored from 0 to 10 points, were appraised by patients themselves. Psychological statuses were measured by 5 mental problems: (A) little interest or pleasure in doing things; (B) felling down, depressed of hopeless; (C) feeling nervous, anxious or on edge; (D) not being able to stop or control worrying; and (E) sleep disorders. Five response options ranged from “not at all” to “nearly every day” to describe the frequency of each item in the previous 2 weeks. Local prevalence of COVID-19 and quarantine measures were also inquired.

### Statistical analysis

Data analysis was conducted using SPSS software, version 24 (SPSS Inc, Chicago, Illinois, USA). The data are presented as the mean ± SD for normally distributed continuous variables, the median and range for skewed-distributed continuous variables, or the number and proportion (%) for categorical variables. Data were compared between groups using the Fisher’s exact test or chi-square test for categorical variables and the Student t test or Mann-Whitney u test for continuous variables. A p value <0.05 was considered to indicate statistically significant differences.

## Results

### Characteristics of the study population

Of 182 questionnaires distributed, 158 were returned (response rate: 86.8%). In total, 157 valid questionnaires rechecked for completeness awere retrieved in our study. The mean age of these patients was 38.4±12.3 years, and 105 patients (66.9%) were females. Among all 157 participants, 156 were not infected with COVID-19 nor close contacts. Only one patient with contact history was isolated strictly for a period and was finally confirmed to be not infected. Eighteen (11.5%) patients had a disease duration less than 1 year, 93 (59.2%) 1 to 5 years, and 46 (29.3%) more than 5 years. A total of 95 (60.5%) patients reported that they had skin lesions, and 57 (36.3%) patients had nail manifestations, while 125 (79.6%) patients had bone lesions with a mean VAS score of 3.3±2.0. In terms of disease activity, the disease aggravated, improved, recurred, and stabilized in 18 (11.5%), 32 (20.4%), 22 (14.0%), and 85 (54.1%) patients, respectively.

### The effects on SAPHO syndrome therapy during COVID-19 epidemic

Therapeutic medication used during the epidemic was variable, including non-steroidal anti-inflammatory drugs (NSAIDs), disease modifying anti-rheumatic drugs (DMARDs) or glucocorticosteroids (47, 29.9%), biologicals (27, 17.2%), traditional Chinese medicine (111, 70.7%), and drugs for external use (11, 7.0%). (Table 1) More than half of the participants (90, 57.3%) claimed that they took medicine regularly, while 67 (42.7%) had medication withdrawal or dose reduction for lack of drugs (26/67, 38.8%), irregular daily schedule (11/67, 16.4%) and subjective reasons (30/67, 44.8%). The duration of irregular treatment varied, including less than 1 week (13, 23.2%), 1–2 weeks (7, 12.5%), 2–4 weeks (9, 16.1%), and more than 4 weeks (27, 48.2%) (Table 2).

**Table 1.** Demographic and clinical characteristics of patients with SAPHO

| Variables |  |
| --- | --- |
| Age (years), mean (SD) | 38.4 (12.3) |
| Sex (Males/Females), n (%) | 52/105 (33.1/66.9) |
| Disease duration |  |
| Less than 1 year, n (%) | 18 (11.5) |
| 1-5 years, n (%) | 93 (59.2) |
| More than 5 years, n (%) | 46 (29.3) |
| Skin lesions, n (%) | 95 (60.5) |
| Nail involvement, n (%) | 57 (36.3) |
| Bone lesions, n (%) | 125 (79.6) |
| VAS, mean (SD) | 3.3 (2.0) |
| Disease activity |  |
| Aggravated, n (%) | 18 (11.5) |
| Improved, n (%) | 32 (20.4) |
| Recurred, n (%) | 22 (14.0) |
| Stable, n (%) | 85 (54.1) |
| Therapeutic drugs |  |
| NSAIDs, DMARDs or glucocorticosteroids, n (%) | 47 (29.9) |
| Traditional Chinese medicine, n (%) | 111 (70.7) |
| Biologicals, n (%) | 27 (17.2) |
| Drugs for external use, n (%) | 11 (7.0) |

SD, standard deviation; NSAID, non-steroidal anti-inflammatory drugs; DMARDs, bisphosphonates, glucocorticosteroids, and disease modifying anti-rheumatic drugs.

**Table 2.** Life and mental conditions of the patients during COVID-19 epidemic

| Variables |  |
| --- | --- |
| Psychologic stress self-assessment, mean (SD) | 3.0 (2.4) |
| Worried about disease condition, n (%) | 65 (41.4) |
| Sources of worries |  |
| Disease aggravation, n (%) | 40 (61.5) |
| Delay of further consultation, n (%) | 48 (73.8) |
| Drug shortage, n (%) | 30 (46.2) |
| Problems of daily life, n (%) | 15 (23.1) |
| Drug withdrawal or dose reducing, n (%) | 67 (42.7) |
| Duration of drug withdrawal or dose reducing |  |
| Less than 1 week, n (%) | 13 (23.2) |
| 1-2 weeks, n (%) | 7 (12.5) |
| 2-4 weeks, n (%) | 9 (16.1) |
| More than 4 weeks, n (%) | 27 (48.2) |

SD, standard deviation.

### Psychological statuses assessment in patients with SAPHO syndrome

Self-assessed psychologic stress score had a mean of 3.0 ± 2.4. A total of 65 (41.4%) patients reported that they worried about their disease conditions because of the epidemic situation. Patients’ concern including disease aggravation (61.5%), delay of further consultation (46.2%), medication shortage(73.8%), and problems of daily life(23.1%) (Table 2). In addition, 78 (49.7%) participants presented that inconvenience in daily life resulted from the epidemic situation troubled them most. And the number of people who had been able to fit in or adapted well to the situation was 92 (58.6%).

Frequency of patients who had been troubled by psychological problems in previous 2 weeks was shown in figure 1. The occurance rates of those problems were in the following order: little interest or pleasure in doing things (104, 66.2%), feeling nervous, anxious, or on edge (90, 57.3%), feeling down, depressed or hopeless (81, 51.6%), sleep disorders (72, 45.9%), not being able to stop or control worrying (69, 43.9%).

**Figure 1.**
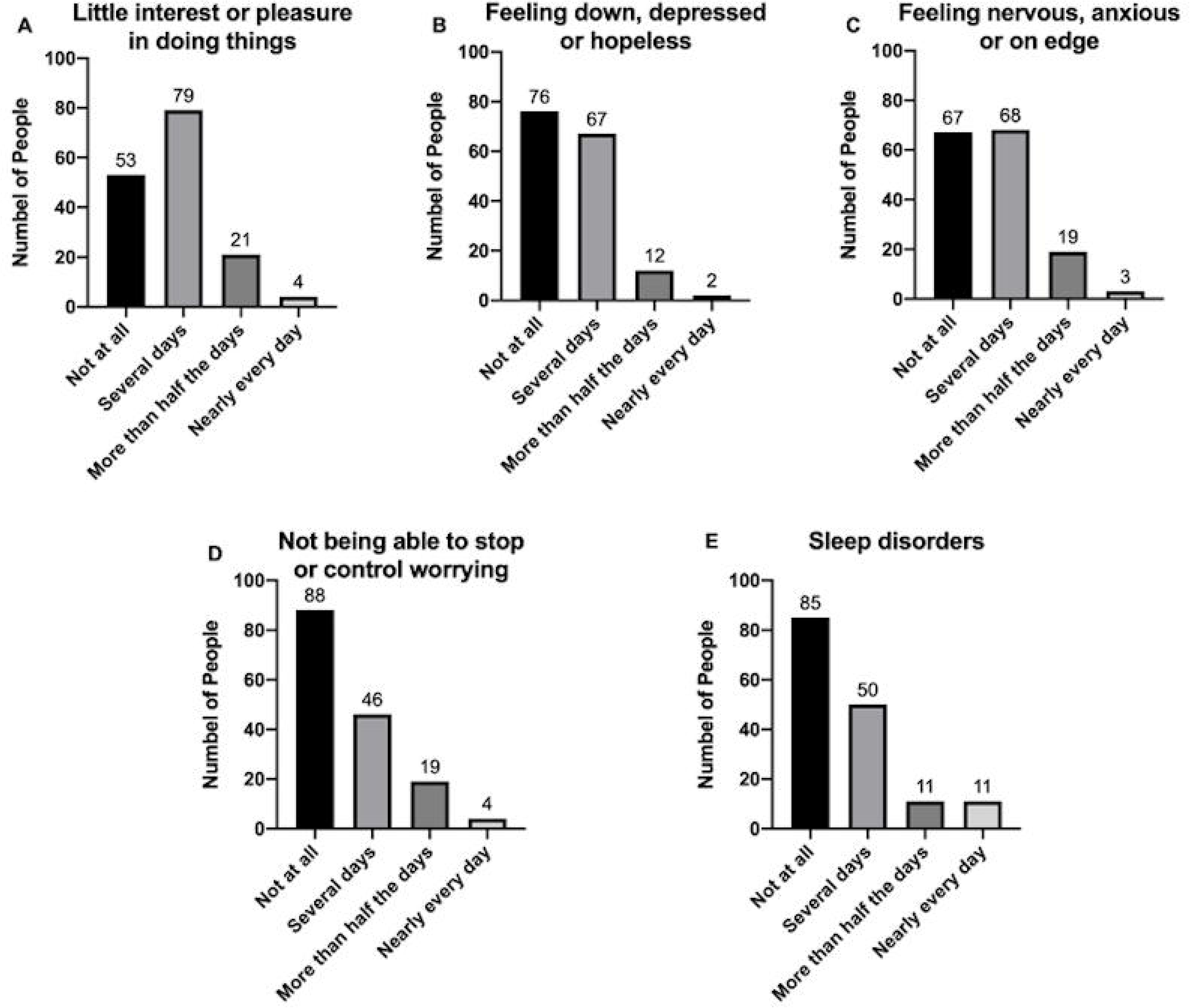
Frequencies of patients troubled by the problems in the past 2 weeks (n=157) (A) little interest or pleasure in doing things; (B) felling down, depressed of hopeless; (C) feeling nervous, anxious or on edge; (D) not being able to stop or control worrying; (E) sleep disorders.

### Effect factors of SAPHO patients’ psychologic situations

Generally, patients with higher disease activity had higher mental pressure. Between VAS score of bone lesions and self-assessed stress score, there exists a significant positive correlation (r=0.418, p<0.0001), which demonstrated that bone and arthrosis pain was associated with negative emotions.

According to their reported disease conditions, participants were divided into three groups which were recurred or aggravated group, stable group, and improved group. There was a significant higher percentage of the worry about the disease conditions because of epidemic situation in the recurred or aggravated group than in the stable group (67.5% vs 37.6%, p=0.002). Similar condition could be found in analysis between the recurred or aggravated group and the improved group (67.5% vs 18.8%, p<0.001). But the stable group and the improved group show no distinct difference on this issue (37.6% vs 18.8%, p=0.040). (Figure 2) Patients with progressive disease condition were more disturbed by inconvenience situation during the epidemic.

**Figure 2.**
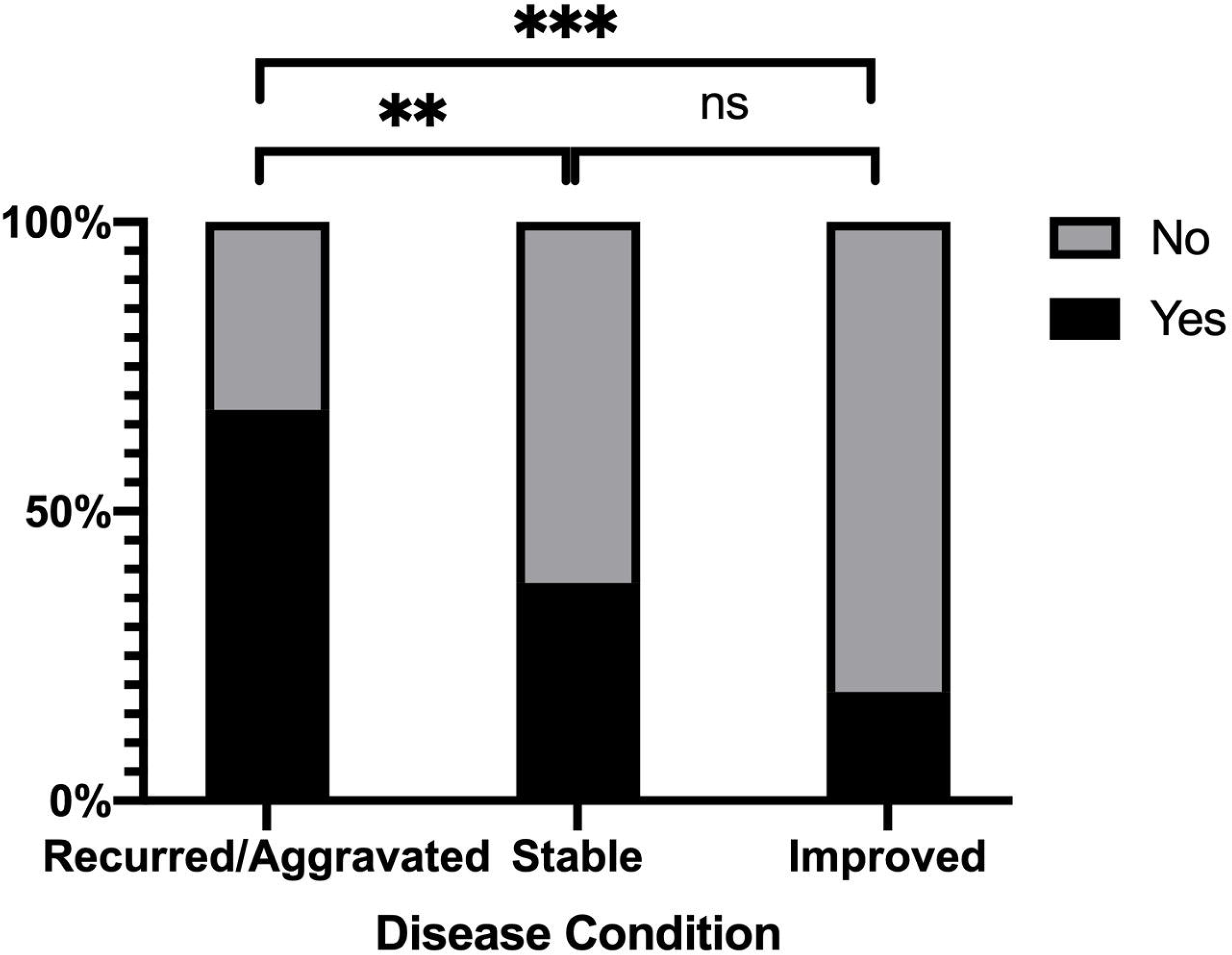
Proportion of patients worried because of epidemic situation with different disease conditions. (\*\**p*<0.01, \*\*\**p*<0.001, ns: not significant)

Different disease manifestations also had different influence on SAPHO patients’ mentalities.A greater percentage of patients with nail involvement felt worried about their disease conditions than patients without (59.6% vs 31.0%, p=0.001). Occurrence rates of feeling nervous, anxious, or on edge (73.7% vs 48.0%, p=0.002) and having little interest or pleasure in doing things (78.9% vs 59.0%, p=0.011) both showed significant differences between patients with nail involvement and those without. Patients with nail involvement felt nervous, anxious, or on edge and had little interest or pleasure in doing things more frequently. (Table 3)

**Table 3.**
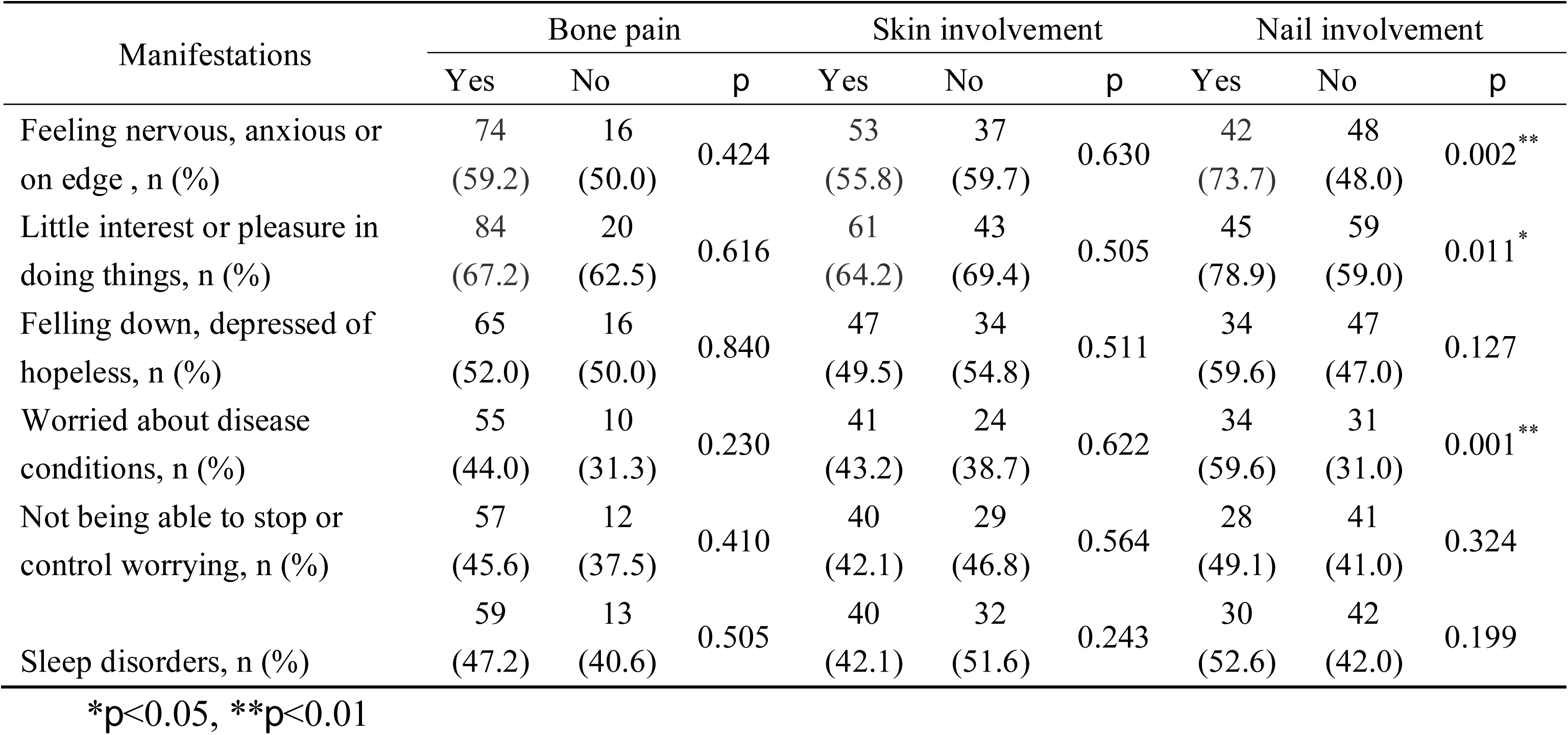
Influence of disease manifestations on SAPHO patients’ mentalities

## Discussion

We presented a detailed analysis of psychological stress of SAPHO patients during the COVID-19 epidemic under different disease conditions and disease manifestations. The COVID-19 epidemic represents a health emergency that is inevitably affecting patients with SAPHO syndrome.

The data showed that the epidemic had a negative psychological effect on SAPHO patients and led to their worries about disease conditions. Owing to restrictions on personal movement, quarantine measures, and fear for COVID-19 infection in the hospital, the majority of patients possibly did not visit the hospital for regular follow-up. Patients with aggravated or recurred diseases might fail to obtain doctors’ advices and help to adjust their treatment plan in time. Consequently, they were more worried about their own disease conditions in the COVID-19 epidemic, which was demonstrated by our data. In addition, a lack of medicine caused by restrictions on movement made patients unable to take medicine regularly and cannot maintain effective treatment. It might lead to a decline in their physical health, which further increased psychological pressure and generated negative emotions.

The current research on the psychological effect of COVID-19 mostly focused on frontline health professionals. A recent study revealed that more than half (50.7%) of frontline health professionals reported depression symptoms, besides anxiety (44.7%) and sleep disorders (36.1%).^10^ There are few studies on psychological situations of patients with chronic diseases during the epidemic. A study on patients treated with hemodialysis during Korean MERS-COV outbreak in 2015 showed that the psychophysical stress indicators of patients increased significantly after 2-week isolation, indicating that the epidemic and quarantine measures would have a psychological impact on patients with chronic diseases.^11^ Our result proved that SAPHO patients felt more bored during the epidemic, which might imply the existence of a certain degree of depression. Strict quarantine measures made the public more likely to feel bored, anxious and nervous.

The mental health and quality of life of SAPHO patients have always been concerned. Misdiagnosis, delayed diagnosis, and chronic injury bring great psychological pressure and life burden to patients. In particular, musculoskeletal manifestations significantly aggravates the disease burden.^12^ Our study also demonstrated the positive correlation between the psychological stress score and VAS score. Therefore, treatment for bone involvement can be beneficial to both of the patients’ physical and mental health. This also confirmed the necessity of sufficient treatment and regular follow-up in patients with SAPHO syndrome during the COVID-19 epidemic.

In addition, there were concerns that whether the use of immunosuppressants will increase the risk of COVID-19 infection in patients with chronic autoimmune diseases. For SAPHO syndrome, NSAIDs are used as first-line treatment for pain relief but always be insufficient. Glucocorticoids, DMARDs, diphosphonates, and biologic DMARDS (eg. anti-TNF-α agents and anti-IL-1 agents) also show their potential efficacy in previous studies.^13–15^ Due to lack of SARS-CoV-2 patients in our survey, we have no evidence to prove the relevance between immunity inhibitors and the risk of COVID-19 infection. Studies on inflammatory bowel diseases^16^ and rheumatoid arthritis^17^ showed that treatment for these diseases might not increase the risk of COVID-19, as uncontrolled disease activity was much riskier. Thus, patients were encouraged to keep on regular treatment.

Our results indicated that SAPHO patients with nail involvement were more likely to be worried and anxious. Nail involvement in SAPHO syndrome is part of clinical manifestations of palmoplantar pustulosis. It could be found in around 30% PPP patients in the form of subungual pustules, onycholysis, pitting, destruction of the nail, or discoloration.^18^ Nail involvement might refer to uncontrolled inflammation and predict more severe skin symptoms^19^, inducing a greater burden of the patient’s life and psychology. It was pointed out that patients with fingernail psoriasis may have more social and psychological burden due to nail pain ^20^ and concerns about the others’ perspective on their nails.^21^ There were more anxiety or depression in patients with psoriasis and nail involvement than those only with skin lesions.^22^ Nail involvement hindered patients’ daily life and work, which led to psychological problems and a decline in quality of life. Thus, in SAPHO syndrome, psychological situations of nail involvement patients should be concerned.

The psychological problems in the public caused by COVID-19 have aroused attention of related government departments. On January 27^th^, 2020, Centre for Disease Prevention and Control of China issued the “Principles of Emergency Psychological Crisis Intervention for the COVID-19 Pneumonia Epidemic”,^23^ which divided the target population into 4 levels. As a group affected by the epidemic prevention and control measures, chronic disease patients are classified as level 4 population together with susceptible people and the public. However, considering the double impact of the epidemic on the physical and mental health of chronic disease patients, it is suggested that more attention should be paid to their psychological and medical care.

According to this survey, we proposed some suggestions on the psychological and medical care of patients with SAPHO syndrome. First of all, pay attention to patients’ psychological and disease conditions. For patients with aggravated or relapsed disease, provide available psychological support and guidance. Psychological problems as stress and anxiety of patients with severe ostealgia or nail involvement should be noted. In addition, ensure timely follow-up visits and adequate medication supply by online consultant and medication distribution. Timely and effective treatment can help relieve patients’ pain, thereby improving their psychological conditions. Additionally, psychological intervention would help reduce the incidence of post-traumatic stress disorder, depression and anxiety.^24,25^

China has now widely launched online psychological intervention services, including psychological counseling via telephone hotlines and intervention platforms.^26^ Considering that the current epidemic of Covid-19 has spread worldwide, the psychological problems of the public might persist or further develop for a long period. It would remain a problem worthy of consideration and attention that how to better allocate limited medical resources to provide psychophysiological support for patients with chronic diseases and rare diseases, such as SAPHO syndrome.

Due to research design, our survey has several limitations. Firstly, only the subjective score was used to quantify the psychological stress. Secondly, their current disease situations were reported subjectively by patients due to the fact that patients cannot be revisited in time with restricted movement. Nonetheless, in the analysis, relatively objective problems were adopted as much as possible to ensure the accuracy and validity of the data in psychological and disease assessment, such as the frequency of a certain psychological problem, whether be worried about the disease condition for the epidemic, and organ involvement.

## Conclusion

The COVID-19 epidemic has a negative effect on the mentality of SAPHO patients. Patients with high disease activity (relapsed or aggravated), severe bone pain, and nail involvement would be more stressed. Physicians and health policy makers should collaborate to provide high-quality, timely psychological services and chronic disease management to vulnerable population like patients with SAPHO syndrome during the COVID-19 epidemic.

## Data Availability

The datasets used or analysed during the current study are available from the corresponding author on reasonable request.

## Acknowledgments

The authors apologize to all colleagues whose works have not been separately cited or discussed here due to space or knowledge limitations. This work was supported by the CAMS Innovation Fund for Medical Sciences [2017-I2M-3–001]; the Capital Medical Research and Development Fund [2016–4–40112]; and the National Key Research and Development Program of China [2016YFC0901500].

